# Factors associated with severity of COVID-19 disease in a multicenter cohort of people with HIV in the United States, March-December 2020

**DOI:** 10.1101/2021.10.15.21265063

**Authors:** Adrienne E. Shapiro, Rachel A. Bender Ignacio, Bridget M. Whitney, Joseph A. Delaney, Robin M. Nance, Laura Bamford, Darcy Wooten, Jeanne C. Keruly, Greer Burkholder, Sonia Napravnik, Kenneth H. Mayer, Allison R. Webel, H. Nina Kim, Stephen E. Van Rompaey, Katerina Christopoulos, Jeffrey Jacobson, Maile Karris, Davey Smith, Mallory O. Johnson, Amanda Willig, Joseph J. Eron, Peter Hunt, Richard D. Moore, Michael S. Saag, W. Christopher Mathews, Heidi M. Crane, Edward R. Cachay, Mari M. Kitahata, for the CFAR Network of Integrated Clinical Systems

## Abstract

**Background:** Understanding the spectrum of SARS-CoV-2 infection and COVID-19 disease in people with HIV (PWH) is critical to provide clinical guidance and implement risk-reduction strategies.

**Objective:** To characterize COVID-19 in PWH in the United States and identify predictors of disease severity.

**Design:** Observational cohort study.

**Setting:** Geographically diverse clinical sites in the CFAR Network of Integrated Clinical Systems (CNICS)

**Participants:** Adults receiving HIV care through December 31, 2020.

**Measurements:** COVID-19 cases and severity (hospitalization, intensive care, death).

**Results:** Of 16,056 PWH in care, 649 were diagnosed with COVID-19 between March-December 2020. Case fatality was 2%; 106 (16.3%) were hospitalized and 12 died. PWH with current CD4 count <350 cells/mm^3^ (aRR 2.68; 95%CI 1.93-3.71; P<.001) or lowest recorded CD4 count <200 (aRR 1.67; 95%CI 1.18-2.36; P<.005) had greater risk of hospitalization. HIV viral load suppression and antiretroviral therapy (ART) status were not associated with hospitalization, although the majority of PWH were suppressed (86%). Black PWH were 51% more likely to be hospitalized with COVID-19 compared to other racial/ethnic groups (aRR 1.51; 95%CI 1.04-2.19, P=.03). Chronic kidney disease (CKD), chronic obstructive pulmonary disease, diabetes, hypertension, obesity, and increased cardiovascular and hepatic fibrosis risk scores were associated with higher risk of hospitalization. PWH who were older, not on ART, with current CD4 <350, diabetes, and CKD were overrepresented amongst PWH who required intubation or died.

**Limitations:** Unable to compare directly to persons without HIV; underestimate of total COVID-19 cases.

**Conclusions:** PWH with CD4 <350 cells/mm^3^, low CD4/CD8 ratio, and history of CD4 <200, have a clear excess risk of severe COVID-19, after accounting for comorbidities also associated with severe outcomes. PWH with these risk factors should be prioritized for COVID-19 vaccination, early treatment, and monitored closely for worsening illness.

## Introduction

The COVID-19 pandemic has had profound direct and structural effects on the health of people with HIV (PWH) in the United States.^1^ The first observations of COVID-19 in PWH occurred during a period of hospital crowding and rationed testing and treatment^2-4^. A large population-based study from South Africa was the first to indicate a two-fold higher risk of COVID-19 mortality among PWH^5^, but this study had limited ability to evaluate the contribution of comorbid conditions. Subsequent global registry data also showed elevated COVID-19 mortality risk associated with HIV, but the contribution of HIV-specific immune compromise and comorbidities were difficult to assess in these studies^6, 7^.

Understanding HIV-associated risks for poor COVID-19 outcomes is important since PWH are often already marginalized and experience health disparities driven in part by social determinants of health. These factors increase the risk of both exposure to COVID-19 and greater COVID-19 severity^8^. PWH experience a disproportionate burden of medical comorbidities^9^, higher rates of smoking^10^, drug and alcohol use^11-15^, and socioeconomic and racial disparities^16,17^. Therefore, disentangling the direct effects of HIV on COVID-19 outcomes is complex, even after adjustment for comorbidities and demographic characteristics. Not all PWH have overt immunosuppression, but HIV itself, chronic immune activation and exhaustion, and metabolic complications of HIV and antiretroviral therapy (ART) contribute to non-AIDS-defining morbidity, even in PWH with high CD4 counts and suppressed viral load (VL)^18-20^. All of these factors could modulate the risk of COVID-19 severity. Understanding the impact of HIV-associated factors on COVID-19 progression remains important to enable clinicians to provide appropriate risk assessment and prioritize COVID-19 prevention and treatment efforts, especially in settings where such resources remain limited.

The objective of this study was to identify predictors of COVID-19 severity, including risk for hospitalization, need for mechanical respiratory support, and death, among PWH who were diagnosed with COVID-19 in the US.

## Methods

The Centers for AIDS Research (CFAR) Network of Integrated Clinic Systems (CNICS) is a prospective observational cohort study of adult PWH in clinical care at academic institutions across the United States^21^. The study cohort included all PWH in care defined as those who attended one or more in-person or virtual HIV primary care visits between September 1, 2018 and December 31, 2020 at seven CNICS sites: Johns Hopkins University, Case Western Reserve University, Fenway Health, University of Alabama at Birmingham, University of California-San Diego, University of North Carolina at Chapel Hill, and University of Washington. CNICS research has been approved by the institutional review boards at each site.

Methods of data collection in CNICS are previously reported^21^. Briefly, comprehensive clinical data including diagnoses, laboratory test results, and medications collected through electronic medical records and institutional data systems undergo rigorous quality assessment and are harmonized in a central repository that is updated quarterly. Demographic data including birth sex and patient-reported racial/ethnic identity were collected at cohort enrollment. Patient Reported Outcome (PRO) measures of smoking (current vs. former vs. never tobacco use) and unstable housing/homelessness were collected through tablet-based surveys every 4-6 months during primary care visits^22, 23^, and from medical records.

### SARS-CoV-2 infections and Outcomes

Candidate SARS-CoV-2 infections and COVID-19 cases were identified through laboratory test results and provider documented diagnoses (ICD-10 codes) recorded between March 1 and December 31, 2020 and verified through medical record review. A positive SARS-CoV-2 RT-PCR or antigen test was considered a verified case. Medical records for all cases identified by diagnosis only without laboratory verification were reviewed by site clinicians and verified as clinical diagnoses using a standardized protocol based on one or more of: 1) a documented report of a positive laboratory test in the chart (e.g. patient had a COVID-19 test result not appearing in the electronic medical laboratory records) or 2) a compatible clinical syndrome and epidemiologic setting (e.g., documented contact), with or without a subsequent positive antibody test result. SARS-CoV-2 results and medical records, including hospitalizations, available to CNICS sites from external health systems were reviewed.

Disease severity indicated by hospitalization for COVID-19, requirement for supplemental oxygen, intensive care admission, and invasive mechanical ventilation were verified by site clinicians and central clinician review of hospitalization discharge summaries. A patient with a positive SARS-CoV-2 test obtained while hospitalized for reasons unrelated to COVID-19 (e.g. pre-operative screening) was considered a verified case but not as hospitalized due to COVID-19. Deaths occurring within 30 days of COVID-19 diagnosis or discharge from a hospitalization due to COVID-19 were attributed to COVID-19.

### Covariates

We examined the following chronic comorbid conditions: diabetes defined using a previously validated approach as: hemoglobin A1c (HbA_1c_) ≥6.5%, a prescription of a diabetes-specific medication, or a diagnosis of diabetes with associated prescription^24^; treated hypertension as a diagnosis of hypertension and a prescription of an anti-hypertensive medication; obesity as body mass index (BMI) ≥30 kg/m^2^; hepatitis C virus (HCV) coinfection by the presence of positive HCV antibody or detectable RNA or genotype; and chronic obstructive pulmonary disease (COPD) using a previously validated approach^25^ as a diagnosis of COPD and ≥90-day continuous supply of long-acting controller medications. We used laboratory test data to compute clinical measures of chronic kidney disease (CKD) defined as last glomerular filtration rate (eGFR) <60 using CKD-EPI without race adjustment^30, 31^, and risk scores for atherosclerotic cardiovascular disease (ASCVD^26^) and hepatic fibrosis (Fibrosis-4: FIB-4^27-29^). We examined CD4 counts (cells/μL) as both lowest historical value, as a proxy for CD4 nadir, and current CD4 count, as well as HIV VL (copies/mL). All laboratory test results were censored one week prior to COVID-19 diagnosis to avoid confounding.

### Statistical analysis

Relative risks for COVID-related hospitalization were calculated using relative risk regression^32^. We accounted for confounding by adjusting models with disease risk scores (DRS)^33, 34^, a prognostic analogue of propensity scores^35^ useful when studying a limited number of exposed patients and outcomes and a relatively large number of confounders. DRS were constructed independently for each exposure of interest using logistic regression in the full cohort^34^ for all non-duplicative covariates (e.g., FIB-4 not adjusted for age, as age is a component of FIB-4 calculation) including age, birth sex, race/ethnicity, smoking status, diabetes, hypertension, and CNICS site. We conducted sensitivity analyses adding comorbid conditions to the DRS to further evaluate the specific effects of race/ethnicity and age. All analyses were conducted in Stata version 17 (StataCorp, College Station, TX).

## Results

Of the 16,056 PWH in care, 649 were identified as COVID-19 cases between March and December 2020; 468 (72%) had documented positive SARS-CoV-2 RT-PCR or antigen test results in the electronic laboratory record, 149 (23%) had a verified COVID-19 diagnosis (including cases with positive PCR tests not appearing in the electronic laboratory record), and the remaining 32 (5%) had provider-reported external verified COVID-19 diagnosis and/or laboratory results. Compared with the study cohort of all PWH in care, PWH diagnosed with COVID-19 were 28% female vs. 21% overall, and 52% were age 50 or older vs. median age of 51 (IQR 39-58) overall. Among the 649 PWH diagnosed with COVID-19, 51% were non-Hispanic Black vs. 45% overall, 25% were non-Hispanic White vs. 38% overall, and 20% were Hispanic of any racial identity vs. 13% overall. As in the overall cohort, the vast majority of PWH diagnosed with COVID-19 were on ART (95%) and had an undetectable VL (86%). While 45% of PWH diagnosed with COVID-19 had a lowest CD4 count <200 cells/mm^3^, more than half (66%) had a current CD4 count ≥500 cells/mm^3^ (**Table 1**).

**Table 1.**
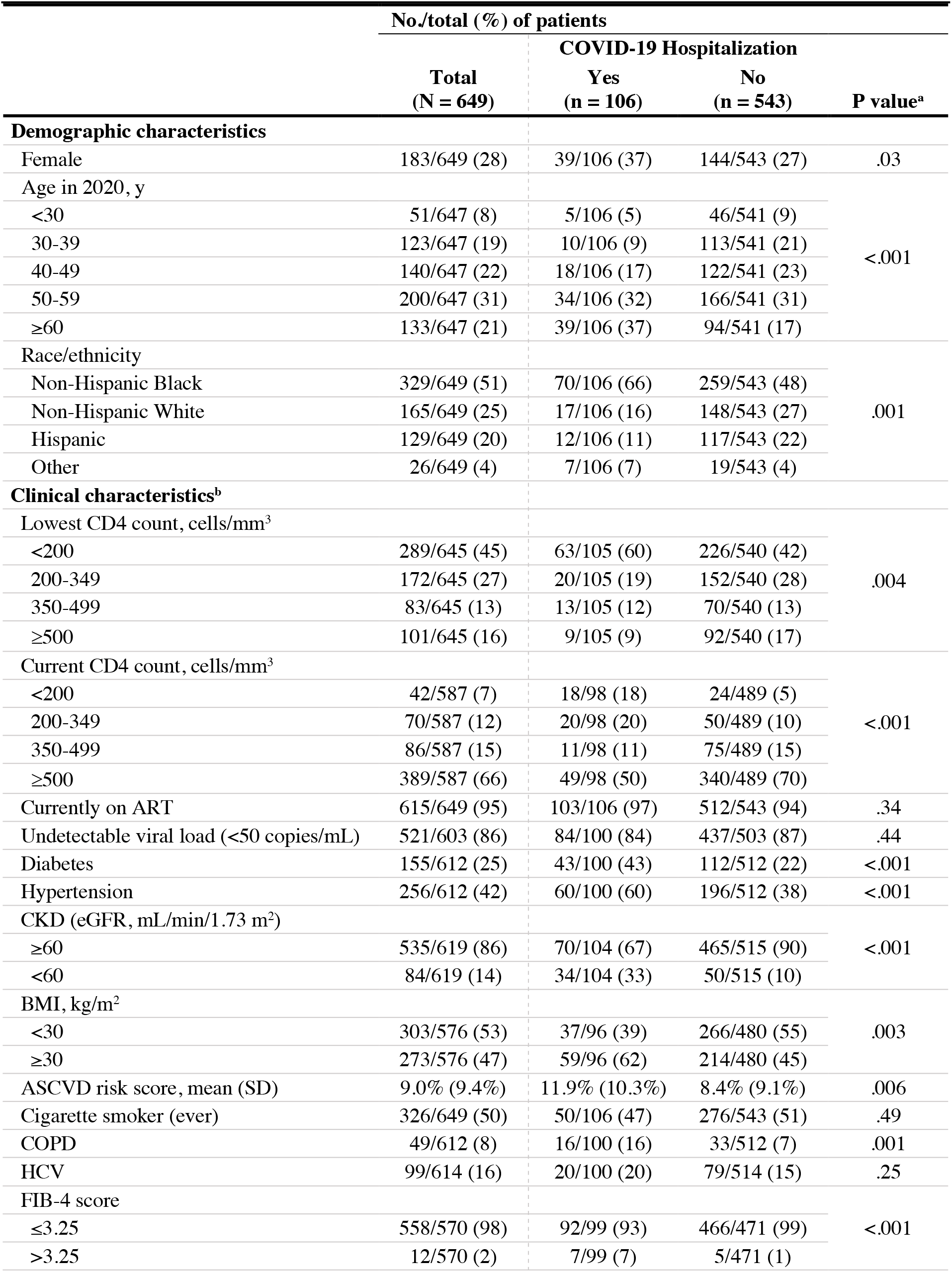
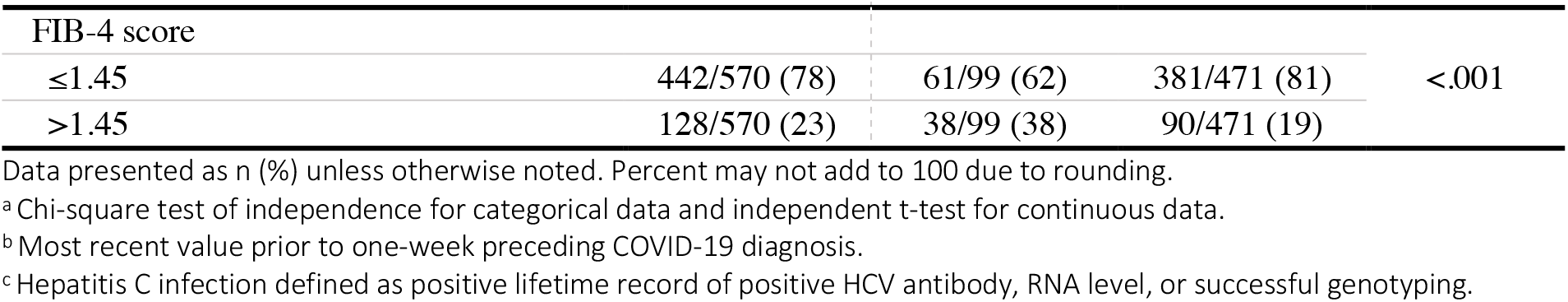
Demographic and clinical characteristics of PWH with COVID-19 by hospitalization status.

One hundred-six (16%) PWH diagnosed with COVID-19 were hospitalized. In univariate analyses, demographic characteristics associated with hospitalization included female sex, Black race, and older age. Compared to non-hospitalized PWH with COVID-19, lowest CD4 count, current CD4 count, elevated FIB-4 and ASCVD risk scores, diabetes, hypertension, CKD (eGFR <60), obesity (BMI ≥30), and COPD, were each associated with risk of COVID-19 hospitalization (all P<.006). Notably, 60% of hospitalized cases had a lowest CD4 count <200 cells/mm^3^ compared with 42% of non-hospitalized cases, and 50% of hospitalized cases had a current CD4 count ≥500 compared with 70% of non-hospitalized cases. Having a FIB-4 score >3.25 (highly predictive of hepatic fibrosis), was strongly associated with hospitalization (7% vs 1%, P<.001). PWH with a FIB-4 score >1.45 were also more likely to be hospitalized with COVID-19 (38% vs 19%, P<.001). HCV coinfection, smoking, and being unstably housed/homeless were not associated with hospitalization among cases with available data.

In adjusted analysis, low current CD4 count was more strongly predictive of hospitalization than age or any comorbid condition. Both current CD4 count <350 aRR 2.68 (95% CI 1.93-3.71; P<.001) and lowest CD4 count <200 aRR 1.67 (95% CI 1.18-2.36; P<.005) were associated with hospitalization (**Figure 1**). There was a trend toward protection against hospitalization with higher current CD4/CD8 ratio, i.e., aRR 0.88 for each 1 SD increase in ratio (95% CI 0.75-1.03; P=.08). There was no association between risk of hospitalization and detectable VL. Although there were insufficient PWH not receiving ART to evaluate an adjusted risk, the proportion of hospitalized cases on ART prior to COVID-19 diagnosis (97%) did not differ significantly from non-hospitalized cases (94%).

**Figure 1.**
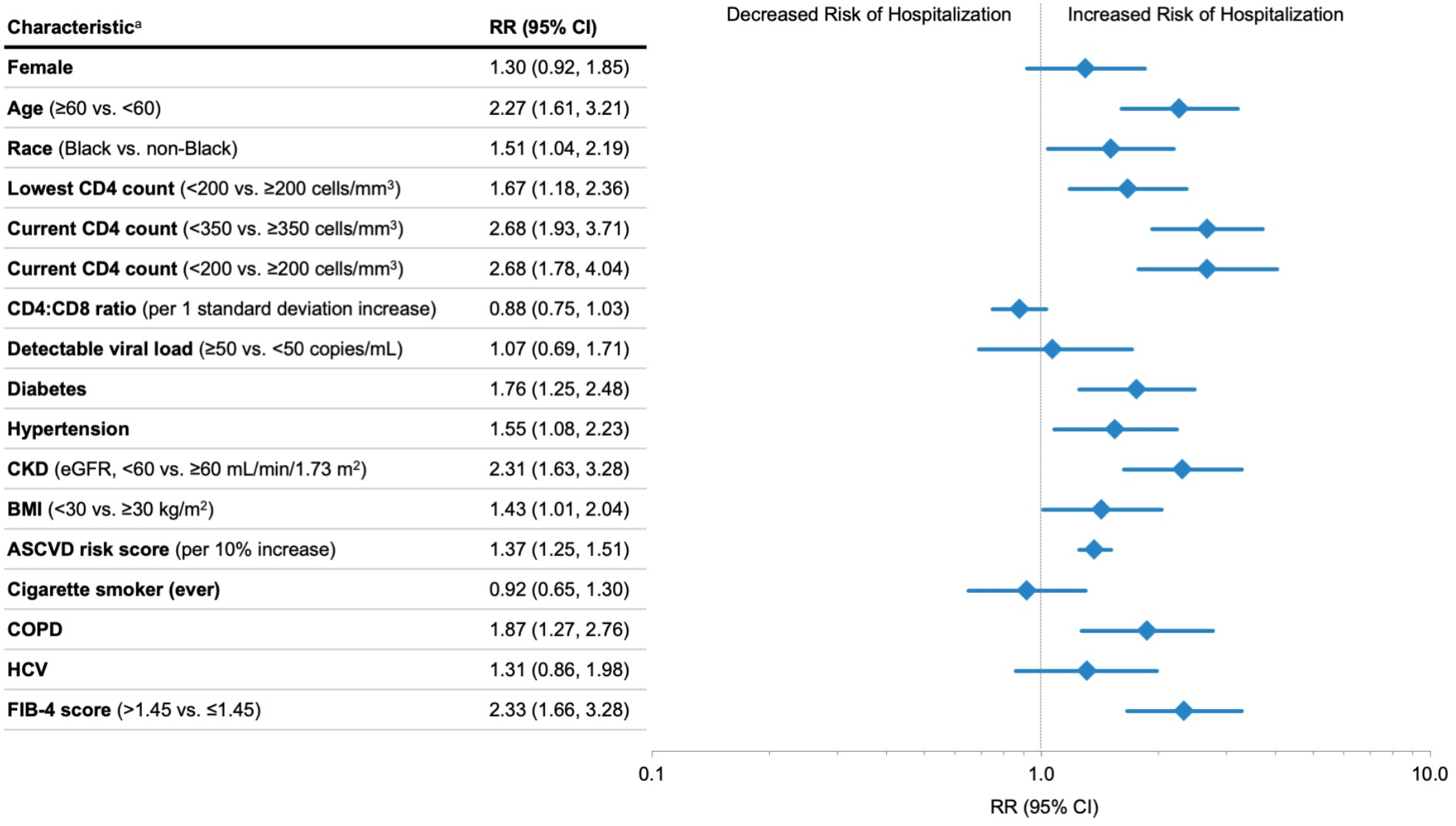
Relative Risk of hospitalization with COVID-19 among PWH with COVID-19 by key characteristics. Abbreviations: ASCVD, atherosclerotic cardiovascular disease; BMI, body mass index; CKD, chronic kidney disease; CNICS, Centers for AIDS Research (CFAR) Network of Integrated Clinical Systems; COPD, chronic obstructive pulmonary disease; eGFR, estimated glomerular filtration rate; FIB-4, Fibrosis-4 scoring system for liver fibrosis; HCV, Hepatitis C virus; PWH, people with HIV; RR, relative risk. ^a^ Relative risk regression models adjusted for demographic and clinical characteristics using disease risk scores, except for the ASCVD risk score analysis which is unadjusted. Disease risk score were constructed independently for each exposure variable of interest using all non-duplicative covariates.

In adjusted analyses, PWH with CKD (eGFR <60) had a more than 2-fold risk of hospitalization (aRR 2.31; 95% CI 1.63-3.28, P<.001), and those with diabetes, hypertension, obesity or COPD also had higher risk of hospitalization (**Figure 1**). In addition, we found a strong association between clinically validated risk scores for cardiovascular and liver disease (ASCVD, FIB-4) and higher risk of hospitalization. For each 10% increase in ASCVD risk score, the aRR for hospitalization was 1.37 (95% CI 1.25-1.51; P<.001). PWH who had a FIB-4 score above the 1.45 cutoff were 2.33 times more likely to be hospitalized with COVID-19, compared to those below this cutoff (95% CI 1.66-3.28; P<.001).

Although women were over-represented among those hospitalized, after adjusting for other factors, sex was not associated with hospitalization (aRR 1.30; 95% CI 0.92-1.85, P=.14). As seen in the general population, age ≥60 was associated with a more than 2-fold risk of hospitalization (aRR 2.27; 95% CI 1.61-3.21, P<.001). PWH identifying as non-Hispanic Black were 51% more likely to be hospitalized with COVID-19 compared to other racial/ethnic identities (aRR 1.51; 95% CI 1.04-2.19, P=.03). Hispanic ethnicity was not significantly associated with hospitalization (aRR=0.71 95% CI 0.41-1.24; P=.23).

To further evaluate whether demographic characteristics associated with COVID-19 hospitalization were partially attributable to the differential distribution of comorbidities, we performed sensitivity analyses that also adjusted for comorbidities including diabetes, hypertension, CKD, obesity, COPD and liver disease (FIB-4), and found similar associations between COVID-19 hospitalization and racial/ethnic identity; the associations between age and hospitalization were attenuated but overall similar.

Twelve PWH diagnosed with COVID-19 (2%) died; three deaths were in persons never hospitalized for COVID-19 and nine died in hospital or within 30 days of discharge. Thirty-one of the 106 hospitalized persons were admitted to the ICU (29%) and 16 (15%) were intubated (**Table 2**). Due to small numbers who were intubated or died, we report proportions but not adjusted risk estimates for critical illness outcomes. Older PWH, those with CD4 count <350, persons not on ART, and those with diabetes and CKD were overrepresented amongst those who required intubation or died from COVID-19. No one under age 30 was admitted to the ICU or died, whereas 25 (81%) of those admitted to the ICU and nine (75%) of the deaths were among PWH 50 and older. As with most COVID-19 hospitalizations in the US, the majority of PWH (65%, **Table 3**) received some degree of oxygen or respiratory support, although presentations with gastrointestinal symptoms, sepsis physiology, acute kidney injury, or COVID-19 pneumonia without significant hypoxemia were observed. Thirty-four percent of hospitalized PWH required only nasal canula as the maximal level of support, 10% required High-Flow Nasal Canula, and 15% were intubated; none received extra-corporeal life support (ECLS/ECMO) **(Table 3)**. While the case fatality rate among PWH with COVID-19 was 12 of 649 (2%), 9% of hospitalized COVID-19 cases died, including nearly one-third (31%) of those intubated. Most non-hospitalized COVID-19 cases had no record of receiving any approved or experimental COVID-19 specific treatment (94%), while 64% of hospitalized COVID-19 cases and 77% of critically ill COVID-19 received COVID-19 pharmacotherapy. Consistent with changes in clinical care during 2020, the most common medications administered to hospitalized PWH with COVID-19 were dexamethasone (45%) and remdesivir (42%), with fewer receiving convalescent plasma or hydroxychloroquine, anti-SARS-CoV-2 monoclonal antibodies, or participating in any interventional trial (**Supplemental Table 1**).

**Table 2.** Demographic and clinical characteristics of PWH hospitalized with COVID-19 by severity of COVID-19 disease.

|  | No./total (%) of patients |  |  |  |
| --- | --- | --- | --- | --- |
|  | Hospitalized<br>(n = 106) | Admitted to ICU<br>(n = 31) | Intubated<br>(n = 16) | Died<br>(n = 12) |
| **Demographic characteristics** |  |  |  |  |
| Female | 39/106 (37) | 12/31 (39) | 5 (31) | 4 (33) |
| Age in 2020, y |  |  |  |  |
| <30 | 5/106 (5) | 0/31 (0) | 0/16 (0) | 0/12 (0) |
| 30-39 | 10/106 (9) | 4/31 (13) | 2/16 (13) | 1/12 (8) |
| 40-49 | 18/106 (17) | 2/31 (7) | 0/16 (0) | 2/12 (17) |
| 50-59 | 34/106 (32) | 9/31 (29) | 4/16 (25) | 2/12 (17) |
| ≥60 | 39/106 (37) | 16/31 (52) | 10/16 (63) | 7/12 (58) |
| Race/ethnicity |  |  |  |  |
| Non-Hispanic Black | 70/106 (66) | 7/31 (23) | 4/16 (25) | 2/12 (17) |
| Non-Hispanic White | 17/106 (16) | 19/31 (61) | 10/16 (63) | 7/12 (58) |
| Hispanic | 12/106 (11) | 3/31 (10) | 1/16 (6) | 3/12 (25) |
| Other | 7/106 (7) | 2/31 (7) | 1/16 (6) | 0/12 (0) |
| **Clinical characteristics<sup>a</sup>** |  |  |  |  |
| Lowest CD4 count, cells/mm<sup>3</sup> |  |  |  |  |
| <200 | 63/105 (60) | 18/31 (58) | 9/16 (56) | 8/12 (67) |
| 200-349 | 20/105 (19) | 4/31 (13) | 2/16 (13) | 1/12 (8) |
| 350-499 | 13/105 (12) | 6/31 (19) | 2/16 (13) | 1/12 (8) |
| ≥500 | 9/105 (9) | 3/31 (10) | 3/16 (19) | 2/12 (17) |
| Current CD4 count, cells/mm<sup>3</sup> |  |  |  |  |
| <200 | 18/98 (18) | 6/26 (23) | 4/15 (27) | 3/11 (27) |
| 200-349 | 20/98 (20) | 5/26 (19) | 2/15 (13) | 3/11 (27) |
| 350-499 | 11/98 (11) | 3/26 (12) | 2/15 (13) | 1/11 (9) |
| ≥500 | 49/98 (50) | 12/26 (46) | 7/15 (47) | 4/11 (36) |
| Currently on ART | 103/106 (97) | 29/31 (94) | 14/16 (88) | 10/12 (83) |
| Undetectable viral load (<50 copies/mL) | 84/100 (84) | 25/29 (86) | 13/15 (87) | 10/12 (83) |
| Diabetes | 43/100 (43) | 15/29 (52) | 9/16 (56) | 6/11 (55) |
| Hypertension | 60/100 (60) | 19/29 (66) | 11/16 (69) | 8/11 (73) |
| CKD (eGFR), mL/min/1.73 m<sup>2</sup> |  |  |  |  |
| ≥60 | 70/104 (67) | 15/30 (50) | 6/16 (38) | 5/12 (42) |
| <60 | 34/104 (33) | 15/30 (50) | 10/16 (63) | 7/12 (58) |
| BMI, kg/m<sup>2</sup> |  |  |  |  |
| <30 | 37/96 (39) | 9/26 (35) | 3/13 (23) | 2/11 (18) |
| ≥30 | 59/96 (62) | 17/26 (65) | 10/13 (77) | 9/11 (82) |
| ASCVD risk score, mean (SD) | 11.9% (10.3%) | 11.7% (10.7%) | 17.2% (13.4%) | 19.6% (12.9%) |
| Cigarette smoker (ever) | 50/106 (47) | 13/31 (50) | 5/16 (31) | 6/12 (50) |
| COPD | 16/100 (16) | 4/29 (14) | 1/16 (6) | 1/11 (9) |
| HCV | 20/100 (20) | 8/29 (28) | 4/16 (25) | 3/11 (27) |
| FIB-4 score |  |  |  |  |
| ≤3.25 | 92/99 (93) | 25/28 (89) | 14/16 (88) | 11/11 (100) |
| >3.25 | 7/99 (7) | 3/28 (11) | 2/16 (13) | 0/11 (0) |

**Table 2. Demographic and clinical characteristics of PWH hospitalized with COVID-19 by severity of COVID-19 disease, continued.**
|  | No./total (%) of patients |  |  |  |
| --- | --- | --- | --- | --- |
|  | Hospitalized<br>(n = 106) | Admitted to ICU<br>(n = 31) | Intubated<br>(n = 16) | Died<br>(n = 12) |
| FIB-4 score |  |  |  |  |
| ≤1.45 | 61/99 (62) | 20/28 (71) | 11/16 (69) | 8/11 (73) |
| >1.45 | 38/99 (38) | 8/28 (29) | 5/16 (31) | 3/11 (27) |

Percent may not sum to 100 due to rounding.
Abbreviations: ART, antiretroviral therapy; ASCVD, atherosclerotic cardiovascular disease; BMI, body mass index; CKD, chronic kidney disease; CNICS, Centers for AIDS Research (CFAR) Network of Integrated Clinical Systems; COPD, chronic obstructive pulmonary disease; eGFR, estimated glomerular filtration rate; FIB-4, Fibrosis-4 scoring system for liver fibrosis; HCV, Hepatitis C virus; PWH, people with HIV.
<sup>a</sup> Most recent value prior to one-week preceding COVID-19 diagnosis.

**Table 3.**
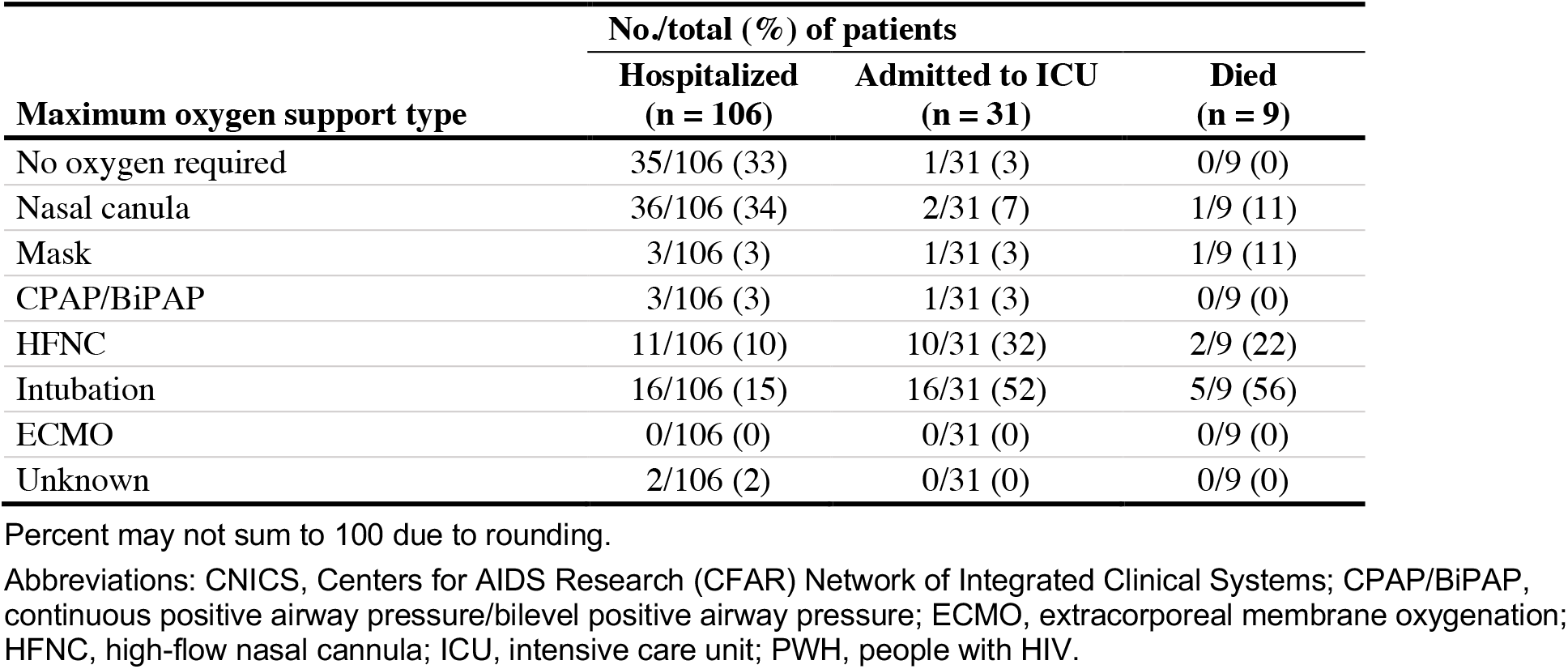
Oxygen requirements among PWH hospitalized with COVID-19 by severity of COVID-19 disease.

| Maximum oxygen support type | No./total (%) of patients |  |  |
| --- | --- | --- | --- |
|  | Hospitalized<br>(n = 106) | Admitted to ICU<br>(n = 31) | Died<br>(n = 9) |
| No oxygen required | 35/106 (33) | 1/31 (3) | 0/9 (0) |
| Nasal canula | 36/106 (34) | 2/31 (7) | 1/9 (11) |
| Mask | 3/106 (3) | 1/31 (3) | 1/9 (11) |
| CPAP/BiPAP | 3/106 (3) | 1/31 (3) | 0/9 (0) |
| HFNC | 11/106 (10) | 10/31 (32) | 2/9 (22) |
| Intubation | 16/106 (15) | 16/31 (52) | 5/9 (56) |
| ECMO | 0/106 (0) | 0/31 (0) | 0/9 (0) |
| Unknown | 2/106 (2) | 0/31 (0) | 0/9 (0) |

Percent may not sum to 100 due to rounding.
Abbreviations: CNICS, Centers for AIDS Research (CFAR) Network of Integrated Clinical Systems; CPAP/BiPAP, continuous positive airway pressure/bilevel positive airway pressure; ECMO, extracorporeal membrane oxygenation; HFNC, high-flow nasal cannula; ICU, intensive care unit; PWH, people with HIV.

## Discussion

We conducted this study in one of the largest and best-characterized multi-center cohorts of PWH and identified factors independently associated with severity of COVID-19 disease in PWH in the US in the first calendar year of the pandemic. Our results demonstrate that both current CD4 count <350 and lowest CD4 <200 cells/mm^3^ are important predictors of hospitalization among PWH with COVID-19. HIV VL suppression was less influential on disease severity, although in our cohort of PWH engaged in care, few were not on ART and suppressed. PWH with CKD and liver dysfunction had over 2-fold higher risk of more severe COVID-19 disease. In addition, PWH with COPD, diabetes, hypertension, and obesity were also at higher risk of greater COVID-19 severity. Using clinically validated and easily measurable risk scores (ASCVD and FIB-4), we found that cardiovascular as well as liver comorbidity were highly predictive indicators of hospitalization for PWH with COVID-19. Our results confirm previously noted associations between comorbidities and COVID-19 severity, but provide more specificity, especially in characterizing the contribution of chronic comorbid conditions and HIV immunologic history to the risk of hospitalization and severity of COVID-19 disease. Although the majority of PWH in our cohort have reconstituted CD4 counts, prior immune destruction or persistent immune activation, as evidenced by low CD4/CD8 ratio, are also important predictors of COVID-19 severity.

Among 16,056 PWH, 106 (0.7%) were hospitalized with COVID-19 between March and December 2020. While identification of non-hospitalized cases was limited by COVID-19 test rationing early in the epidemic, hospitalization with severe disease is less subject to misclassification and bias. Testing for SARS-CoV-2 infection in US hospitals was widely and uniformly available throughout most of the study period, reducing potential for bias in identifying hospitalized patients. The proportions of PWH with COVID-19 requiring hospitalization (16%) and intensive care (5%) and who died (2%) are lower than proportions reported in a Spanish national cohort of PWH with COVID-19 early in the pandemic (64% hospitalized, 6% ICU, 8% died)^36^, and PWH in New York City with COVID-19 in March-June 2020 (42%, 5%, 13%, respectively)^37^, which may be explained in part by overwhelmed healthcare capacity during the limited time period of those early cohort studies. In contrast, PWH in a Spanish (PISCIS) registry through December 2020 and US national registry of COVID-19 patients (NC3 Cohort) through May 2021, had similar rates of severe outcomes (PISCIS: 13.8%, 0.9%, 1.7%; NC3: 32%, 4%, 2%, respectively) to our cohort^38, 39^.

Cardiovascular, pulmonary and metabolic comorbidities in PWH associated with increased disease severity were consistent with those seen in other cohorts of PWH and in people without HIV.^2, 40-42^ Notably, we found that CKD with even modest reductions in eGFR was among the strongest predictors of hospitalization for PWH with COVID, consistent with non-HIV cohorts, and not previously well-recognized in PWH, despite the increased prevalence of CKD in PWH.^40, 41, 43^

Early studies of PWH with COVID-19 did not detect a strong association between CD4 count and severity of COVID-19 outcomes^44, 45^, and initially it was hypothesized that immune suppression may be protective of cytokine-mediated inflammatory responses in COVID-19^46^. Our results demonstrate consistent and significant increased risk of hospitalization in PWH with lower CD4 counts, an association that has been confirmed in multiple global settings^47, 48^. While most marked in PWH with CD4 <200 cells/mm^3^, risk of hospitalization was significantly elevated in people with CD4 <350 compared to CD4 ≥350. In addition to current CD4 count, we also found lowest CD4 count and lower current CD4/CD8 ratio were associated with an increased risk of hospitalization, suggesting an immune exhaustion effect persisting from the time of CD4 nadir despite subsequent CD4 reconstitution. Importantly, in this study, we excluded CD4 counts obtained at or after COVID-19 diagnosis, as SARS-CoV-2 infection can cause profound CD4 and CD8 lymphocyte suppression, which if not carefully parameterized, may have contributed to confounding in other cohort studies. In addition to characterizing HIV-associated risks for hospitalization with greater precision, this study is the first to our knowledge, in PWH or in the general population, to examine ASCVD and FIB-4, which are clinically available, robust indicators of CVD risk and liver comorbidity that were each associated with severity of COVID-19. We recommend these scores be used by clinicians for risk estimation, encouragement of vaccination, and allocation of monoclonal antibodies to prevent disease progression.

Our study also confirmed that minoritized racial and ethnic groups had disproportionately severe COVID-19 outcomes, even within a population predominately representing minoritized groups. Black PWH diagnosed with COVID-19 were 50% more likely to be hospitalized than other PWH, a finding that persisted after controlling for multiple medical comorbidities. Racism, anti-LGBTQ bias, and other forms of discrimination and stigmatization also increase allostatic load, in addition to the structural barriers and health inequities faced by many PWH compounded by marginalized gender identities, sexual orientation, and substance use^16, 49-52^.

The greatest strength of this analysis is the extensive and well-characterized multi-site cohort, in which chronic comorbidities and HIV-specific factors have been previously validated. Our analysis has some limitations. The number of COVID-19 cases identified likely underestimates the true prevalence of SARS-CoV-2 infection and COVID-19 disease. Thus, the proportion of cases who were hospitalized may be an overestimate, though absolute ascertainment of hospitalization and severe outcomes would not be affected. We were unable to make comparisons to the general population; such comparisons have been published, albeit without the rigorous case characterization and clinician hospitalization review as in CNICS.

## Conclusions

Given the experience of many long-term survivors living through the first two decades of the AIDS epidemic, the isolation, scientific uncertainty, and lack of actionable medical interventions has revived many parallel concerns for PWH^17, 53, 54^. Developing clear understanding of whom amongst those with HIV is at increased risk for severe outcomes with this new pathogen, and which risks are modifiable, can mitigate both the medical and psychological burden to PWH during this pandemic. Results of this large multi-site cohort study demonstrate that older PWH with a CD4 cell count <350 or historic CD4 <200 have higher risk of severe COVID-19 disease. Comorbidities including CKD, liver fibrosis, COPD, diabetes, hypertension, obesity and cardiovascular disease also impart increased risk of severe COVID-19 in PWH.

## Data Availability

All data produced in the present work are available from the CNICS Study and are available upon application to the CNICS Investigators. https://sites.uab.edu/cnics/submit-a-proposal/

## Acknowledgments

We gratefully acknowledge the CNICS cohort participants and their providers for contributing to the study. Funding for this study and for CNICS came from the National Institute of Allergy and Infectious Diseases (NIAID) [CNICS R24 AI067039; UW CFAR NIAID Grant P30 AI027757; UAB CFAR grant P30 AI027767; UNC CFAR grant P30 AI50410; UCSD CFAR grant P30 AI036214; Case Western Reserve University CFAR grant P30 AI036219; Fenway Health/Harvard CFAR grant P30 AI060354, UCSF CFAR grant P30 AI027763 and JHU CFAR grant P30 AI094189] and the National Institute on Drug Abuse (NIDA) [R01DA047045].

---

**Supplementary Table 1.**
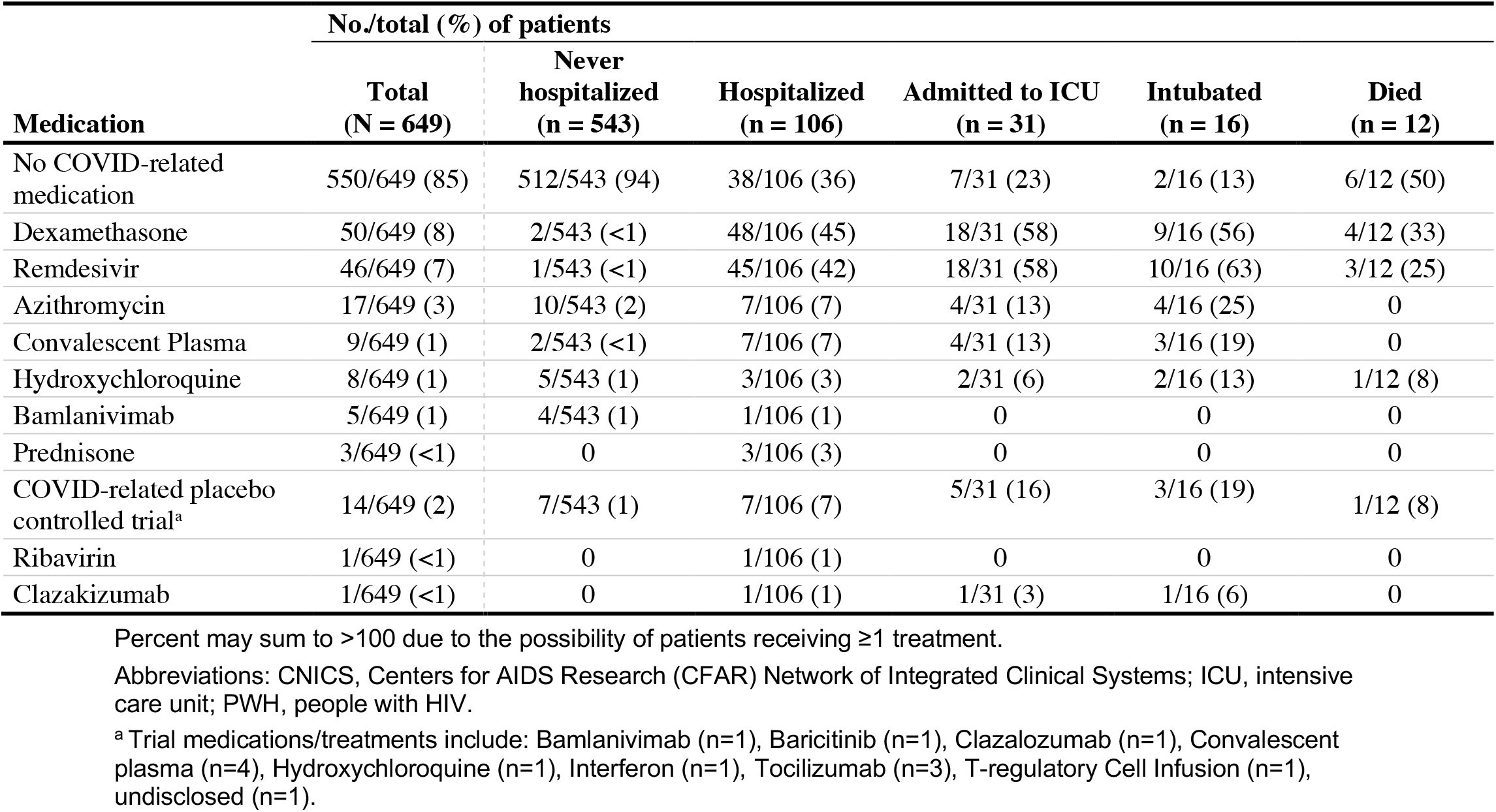
COVID-19 medications among PWH by severity of COVID-19 disease.

