## Supplemental Table 1 for "Factors associated with severity of COVID-19 disease in a multicenter cohort of people with HIV in the United States, March-December 2020"

| **Supplementary Table 1. COVID-19 medications among PWH by severity of COVID-19 disease.** | | | | | | |
| --- | --- | --- | --- | --- | --- | --- |
|  | **No./total (%) of patients** | | | | | |
| **Medication** | **Total**  **(N = 649)** | **Never hospitalized**  **(n = 543)** | **Hospitalized**  **(n = 106)** | **Admitted to ICU**  **(n = 31)** | **Intubated**  **(n = 16)** | **Died**  **(n = 12)** |
| No COVID-related medication | 550/649 (85) | 512/543 (94) | 38/106 (36) | 7/31 (23) | 2/16 (13) | 6/12 (50) |
| Dexamethasone | 50/649 (8) | 2/543 (<1) | 48/106 (45) | 18/31 (58) | 9/16 (56) | 4/12 (33) |
| Remdesivir | 46/649 (7) | 1/543 (<1) | 45/106 (42) | 18/31 (58) | 10/16 (63) | 3/12 (25) |
| Azithromycin | 17/649 (3) | 10/543 (2) | 7/106 (7) | 4/31 (13) | 4/16 (25) | 0 |
| Convalescent Plasma | 9/649 (1) | 2/543 (<1) | 7/106 (7) | 4/31 (13) | 3/16 (19) | 0 |
| Hydroxychloroquine | 8/649 (1) | 5/543 (1) | 3/106 (3) | 2/31 (6) | 2/16 (13) | 1/12 (8) |
| Bamlanivimab | 5/649 (1) | 4/543 (1) | 1/106 (1) | 0 | 0 | 0 |
| Prednisone | 3/649 (<1) | 0 | 3/106 (3) | 0 | 0 | 0 |
| COVID-related placebo controlled trial^a^ | 14/649 (2) | 7/543 (1) | 7/106 (7) | 5/31 (16) | 3/16 (19) | 1/12 (8) |
| Ribavirin | 1/649 (<1) | 0 | 1/106 (1) | 0 | 0 | 0 |
| Clazakizumab | 1/649 (<1) | 0 | 1/106 (1) | 1/31 (3) | 1/16 (6) | 0 |

Percent may sum to >100 due to the possibility of patients receiving ≥1 treatment.

Abbreviations: CNICS, Centers for AIDS Research (CFAR) Network of Integrated Clinical Systems; ICU, intensive care unit; PWH, people with HIV.

^a^ Trial medications/treatments include: Bamlanivimab (n=1), Baricitinib (n=1), Clazalozumab (n=1), Convalescent plasma (n=4), Hydroxychloroquine (n=1), Interferon (n=1), Tocilizumab (n=3), T-regulatory Cell Infusion (n=1), undisclosed (n=1).
